# Case-control study of neuropsychiatric symptoms following COVID-19 hospitalization in 2 academic health systems

**DOI:** 10.1101/2021.07.09.21252353

**Authors:** Victor M. Castro, Jonathan Rosand, Joseph T. Giacino, Thomas H. McCoy, Roy H. Perlis

## Abstract

Neuropsychiatric symptoms may persist following acute COVID-19 illness, but the extent to which these symptoms are specific to COVID-19 has not been established. We utilized electronic health records across 6 hospitals in Massachusetts to characterize cohorts of individuals discharged following admission for COVID-19 between March 2020 and May 2021, and compared them to individuals hospitalized for other indications during this period. Natural language processing was applied to narrative clinical notes to identify neuropsychiatric symptom domains up to 150 days following hospitalization. Among 6,619 individuals hospitalized for COVID-19 drawn from a total of 42,961 hospital discharges, the most commonly documented symptom domains between 31 and 90 days after initial positive test were fatigue (13.4%), mood and anxiety symptoms (11.2%), and impaired cognition (8.0%). In models adjusted for sociodemographic features and hospital course, none of these were significantly more common among COVID-19 patients; indeed, mood and anxiety symptoms were less frequent (adjusted OR 0.72 95% CI 0.64-0.92). Between 91 and 150 days after positivity, most commonly-detected symptoms were fatigue (10.9%), mood and anxiety symptoms (8.2%), and sleep disruption (6.8%), with impaired cognition in 5.8%. Frequency was again similar among non-COVID-19 post-hospital patients, with mood and anxiety symptoms less common (aOR 0.63, 95% CI 0.52-0.75). Neuropsychiatric symptoms were common up to 150 days after initial hospitalization, but occurred at generally similar rates among individuals hospitalized for other indications during the same period. Post-acute sequelae of COVID-19 thus may benefit from standard if less-specific treatments developed for rehabilitation after hospitalization.

**Funding:** R01MH120227, R01MH116270 (Perlis)

## Introduction

For a subset of individuals infected with SARS-CoV-2, symptoms may persist from 30 days to 6 months or more following acute COVID-19 illness. While nearly every organ system may be impacted by such symptoms, symptoms arising from central nervous system pathology may be particularly prominent, even in the absence of evidence of direct SARS-CoV-2 involvement of the nervous system[1]. In the largest study to date, among more than 73,000 individuals diagnosed with but not initially hospitalized for COVID-19 in the Veterans Administration health system, rates of psychiatric and neurocognitive diagnoses, as well as pharmacotherapies used to treat them, were markedly elevated compared to a control cohort of >5 million not diagnosed with COVID-19[2]. A complementary study utilizing claims data for ∼27,000 individuals age 18-65 with a COVID-19 diagnosis in the US similarly found elevated rates of memory complaints, anxiety, and fatigue compared to matched controls[3], as did another US claims data set among more than 200,000 COVID survivors which found elevated rates of anxiety, dementia, and psychotic disorders[4]. These results comport with patient-driven self-reports, such as an app-based study of ∼4,000 individuals in whom fatigue and headaches were the most common symptoms among the 13% with symptoms persisting beyond 28 days[5].

Consideration of hospitalized cohorts may facilitate estimates of risk among individuals with more severe illness. In the first such study, a 6-months follow-up of ∼1700 COVID-19-hospitalized patients in Wuhan, China, found rates of sleep disruption, anxiety, and depression to be markedly elevated, along with fatigue and dyspnea[6]. Elevated rates of brain-related sequelae were also observed in a VA cohort of more than 13,000 individuals hospitalized for COVID-19, matched 1:1 to those hospitalized for influenza[2].

While these and other studies indicate that neuropsychiatric post-acute sequelae of COVID-19 (PASC) are common, what is less clear is how specific such symptoms may be. That is, to what extent might they reflect the consequences of any acute and highly morbid illness, rather than COVID-19 in particular. This distinction is more than semantic: if they are specific to COVID-19, efforts to dissect pathophysiology may be particularly important in understanding how to treat them. On the other hand, if they are not, such efforts may be less useful than simply applying standard approaches to rehabilitation following hospitalization. For example, a recent small study suggested that the pattern of neuropsychiatric deficits immediately following hospitalization was non-specific and resembled that observed in individuals without COVID-19[7]. A large claims study found association between greater COVID-19 severity and risk of sequelae – but did not examine specificity of this effect[4].

In understanding specificity, derivation of appropriate comparator groups is critical. While methods such as propensity-score matching are powerful, they rely on availability of a sufficient number of patients on-support – i.e., similar enough – to enable matching. Moreover, convergent evidence using other health systems and designs can increase confidence in the robustness of prior results, even when earlier studies conducted abundant sensitivity analysis[2]. Therefore, we utilized data from 6 Massachusetts hospitals across 2 academic medical centers to examine post-hospitalization neuropsychiatric symptoms. Our work differs from prior work in two key ways. First, we sought to capture symptoms, not diagnostic codes, recognizing that symptoms may be present even if not associated with a formal diagnosis, and may be less susceptible to Berkson’s bias, a form of selection bias which may lead to overrepresentation of comorbid diagnoses. To this end, we applied natural language processing (NLP) to capture individual symptoms, and examined the overlap between such symptoms and corresponding diagnostic codes. Second, we considered all individuals hospitalized at the same time as COVID-19, adjusting for sociodemographic and clinical features, rather than matching on specific diagnoses such as influenza. This allows us to answer the question, ‘are post-acute sequelae of COVID-19 different from post-hospital sequelae in general?’ In both respects, we leveraged electronic health records to investigate post-acute neuropsychiatric sequelae of COVID-19 in a manner that complements and extends recent large claims or diagnostic code-based studies.[2,4]

## Methods

### Study Design and Cohort Derivation

We utilized a retrospective cohort design that included any individual age 18-99 with documented polymerase chain reaction (PCR) test result, positive or negative, who was admitted emergently from any of 6 Eastern Massachusetts hospitals between March 1,2020 and May 15, 2021. Labor and delivery and other elective admissions were not included in the analysis. We further required that patients are discharged alive within 30 days of test-positivity, to yield a more homogeneous follow-up cohort. Clinical notes, ICD10 diagnostic codes and sociodemographic features were extracted from the Mass General Brigham Research Patient Data Registry (RPDR) [8] and used to generate a datamart [9]. As an aggregate measure of illness burden prior to admission, we calculated a comorbidity index using methods previously described [10]. To characterize inpatient course, intensive care unit admission and use of mechanical ventilation were determined from the enterprise data warehouse.

The Human Research Committee of Mass General-Brigham approved this research protocol, granting a waiver of requirement for informed consent as detailed by 45 CFR 46.116, because only secondary use of data generated by routine clinical care was required.

### Phenotype generation

To generate symptom domains, we applied a simple natural language processing (NLP) strategy that we have successfully applied in abundant prior work examining clinical and biological associations with neuropsychiatric symptoms (see, e.g., McCoy and Barroilhet[11,12]). Two of the authors (RP, VC) manually and iteratively curated token lists (i.e., lists of terms that may represent individual symptoms) using terms expanded from a large self-report survey[5,13] to include common synonyms (Supplemental Table 1), reviewing randomly-selected notes to. Presence of at least one such term, without negation (e.g., “not depressed”, “no evidence of depression”) and exclusive of the family history or patient instruction sections of notes, was considered as presence of a documented symptom or sign. To maximize sensitivity to symptoms, we also identified ICD-10 symptom codes (ICD10: R*) corresponding to each symptom domain and combined them with the NLP data (Supplemental Table 2). Thus, presence of a symptom could reflect either documentation or presence of a code. Full token lists and ICD-10 codes used are provided in Supplemental Materials. For 3 symptoms (anosmia, headache, and fatigue) for which ICD-10 codes are available that correspond to symptoms, we compared sensitivity and specificity of the NLP tokens and ICD-10 codes to ‘gold standard’ manually annotated admissions (Supplemental Table 3).

### Analysis

We examined frequency of at least one symptom in a given domain among SARS-CoV-2 PCR test-positive and negative individuals, and then compared these frequencies using logistic regression, without and then with adjustment for sociodemographic features and characteristics of hospital course. Specifically, models were adjusted for hospital type, age at admission, race, Hispanic ethnicity, public insurance, Charlson comorbidity index, ICU admission and mechanical ventilation. (We do not report coefficients for covariates as our intention was not to estimate risk factors for subsequent symptoms per se). These data were not missing for any individuals. For each neuropsychiatric symptom domain, we examined acute (14 days prior to testing through 30 days after testing), 31-90 day, and 91-150 day presence; individuals were only included in analysis if at least one note or diagnostic code was available in that interval and there was sufficient follow-up for each time period. For each symptom we excluded patients with a prior history of a symptom (occurring 18 months to 14 days prior to COVID index date) so that symptoms reported are *incident* to the acute or post-acute time period.

Analyses utilized R 4.0.1 [14]. No correction for multiple-hypothesis testing was applied, with p<0.05 considered the threshold for statistical significance.

## Results

Table 1 summarizes features of the 6,619 individuals hospitalized with COVID-19, drawn from a total of 42,961 hospital discharges. For COVID-19 cases, median age was 63 (IQR 50-76); they were 47% female, 59% White, 14% Black, and 4% Asian; 24% were Hispanic; and 54% had public insurance. The most commonly documented incident symptom domains between 30 and 90 days after initial positive test among COVID-19 cases were fatigue (13.4%), mood and anxiety symptoms (11.2%), and impaired cognition (8.0%). (Figure 1, top; for description of symptoms documented between 14 days prior to test and 30 days following, see Supplemental Figure 1). Cognitive symptoms, fatigue, and hallucinations were all significantly more frequent in the 30-90 day window among COVID-19 cases in unadjusted regression models, but not in models adjusted for sociodemographic features and hospital course. Headache (adjusted OR 0.84, 95% CI 0.71-0.98), language disturbance (aOR 0.42, 95% CI 0.24-0.68), and mood and anxiety symptoms (aOR 0.72, 95% CI 0.64-0.82) were significantly less common in COVID-19 cases (Table 2). Between 91 and 150 days after positivity, the most common symptoms were fatigue (10.9%), mood and anxiety symptoms (8.2%), and sleep disruption (6.8%), with impaired cognition in 5.8% (Figure 1, bottom). Frequency was similar among non-COVID-19 post-hospital patients in crude and adjusted models, with the exception that mood and anxiety symptoms (aOR 0.63, 95% CI 0.52-0.75) remained less common among post-COVID-19 patients in this interval (Table 3).

**Table 1.** Demographic, hospital course and follow-up comparison between COVID positive and COVID negative admissions.

| Characteristic | COVID Positive<br>Admit, N = 6,619<sup>1</sup> | COVID Negative<br>Admit, N = 36,342<sup>1</sup> | p-value<sup>2</sup> |
| --- | --- | --- | --- |
| **<u>Demographics</u>** |  |  |  |
| **Age at admission** | 63 (50, 76) | 65 (51, 77) | <0.001 |
| **Gender** |  |  | <0.001 |
| Female | 3,132 (47%) | 18,184 (50%) |  |
| Male | 3,487 (53%) | 18,157 (50%) |  |
| Unknown | 0 (0%) | 1 (<0.1%) |  |
| **Race** |  |  | <0.001 |
| Asian | 277 (4.2%) | 1,167 (3.2%) |  |
| Black | 941 (14%) | 3,123 (8.6%) |  |
| Other | 1,055 (16%) | 2,368 (6.5%) |  |
| Unknown | 428 (6.5%) | 1,105 (3.0%) |  |
| White | 3,918 (59%) | 28,579 (79%) |  |
| **Hispanic ethnicity** | 1,575 (24) | 3,360 (9.3) | <0.001 |
| **Public insurance** | 3,588 (54%) | 19,547 (54%) | 0.5 |
| **Homeless patient** | 127 (1.9%) | 715 (2.0%) | 0.8 |
| **Health system PCP** | 3,086 (47%) | 17,495 (48%) | 0.023 |
| **Charlson Comorbidity Index** | 1.85 (2.43) | 2.21 (2.67) | <0.001 |
| **<u>Hospital course</u>** |  |  |  |
| **Hospital Type** |  |  | <0.001 |
| Academic Medical Center | 3,414 (52%) | 19,866 (55%) |  |
| Community Hospital | 3,205 (48%) | 16,476 (45%) |  |
| **Admitted via ED** | 6,483 (98%) | 33,805 (93%) | <0.001 |
| **Admitted via Psych ED** | 66 (1.0%) | 496 (1.4%) | 0.015 |
| **Length of stay (days)** | 5 (3, 8) | 4 (2, 6) | <0.001 |
| **ICU admission** | 839 (13%) | 3,820 (11%) | <0.001 |
| **ICU length of stay (hours)** | 90 (40, 232) | 46 (25, 85) | <0.001 |
| **Oxygen therapy or NIV** | 3,987 (60%) | 14,838 (41%) | <0.001 |
| **Mechanical Ventilation** | 400 (6%) | 1,246 (3%) | <0.001 |
| **<u>Follow-up after discharge</u>** |  |  |  |
| **Total follow-up (days)** | 186 (120, 377) | 222 (123, 310) | <0.001 |
| **Follow-up >= 90 days** | 5,771 (87%) | 30,193 (83%) | <0.001 |
| **Follow-up >= 150 days** | 4,056 (61%) | 25,016 (69%) | <0.001 |
| <sup>1</sup>n (%); Median (IQR); Mean (SD) |  |  |  |
| <sup>2</sup>Pearson's Chi-squared test; Wilcoxon rank sum test; Fisher's exact test |  |  |  |

PCP: Primary care provider; ED: Emergency department; ICU: Intensive care unit; NIV: Non-invasive ventilation

**Table 2.** Unadjusted and adjusted odds of neuropsychiatric post-acute sequelae at 31 to 90 days post-admission

| Symptom | 31 – 90d Post-acute<br>Unadjusted Model |  |  |  | 31 – 90d Post-acute<br>Adjusted Model* |  |  |  |
| --- | --- | --- | --- | --- | --- | --- | --- | --- |
|  | OR | 95% CI<br>Low | 95% CI<br>High | P | Adj OR | Adj 95%<br>CI Low | Adj 95%<br>CI High | Adj p |
| Anosmia | 2.04 | 1.64 | 2.53 | <0.001 | 2.12 | 1.69 | 2.63 | <0.001 |
| Cognition | 0.91 | 0.79 | 1.05 | 0.19 | 0.89 | 0.77 | 1.02 | 0.104 |
| Fatigue | 1.00 | 0.90 | 1.12 | 0.94 | 0.98 | 0.88 | 1.10 | 0.761 |
| Hallucinations | 1.70 | 0.99 | 2.76 | 0.04 | 1.54 | 0.89 | 2.53 | 0.105 |
| Headache | 0.85 | 0.73 | 0.99 | 0.03 | 0.84 | 0.71 | 0.98 | 0.025 |
| Language | 0.40 | 0.23 | 0.65 | <0.001 | 0.42 | 0.24 | 0.68 | 0.001 |
| Memory | 0.84 | 0.62 | 1.10 | 0.22 | 0.86 | 0.64 | 1.14 | 0.314 |
| Mood/anxiety | 0.74 | 0.66 | 0.84 | <0.001 | 0.72 | 0.64 | 0.82 | <0.001 |
| Sleep | 0.92 | 0.80 | 1.06 | 0.26 | 0.93 | 0.81 | 1.08 | 0.346 |

**Table 3.** Unadjusted and adjusted odds of neuropsychiatric post-acute sequelae at 91 to 150 days post-admission

| Symptom | 91 – 150d Post-acute<br>Unadjusted Model |  |  |  | 91 – 150d Post-acute<br>Adjusted Model* |  |  |  |
| --- | --- | --- | --- | --- | --- | --- | --- | --- |
|  | OR | 95% CI<br>Low | 95% CI<br>High | p | Adj OR | Adj 95%<br>CI Low | Adj 95%<br>CI High | Adj p |
| Anosmia | 1.19 | 0.83 | 1.65 | 0.323 | 1.23 | 0.86 | 1.72 | 0.240 |
| Cognition | 0.86 | 0.70 | 1.06 | 0.164 | 0.88 | 0.71 | 1.08 | 0.229 |
| Fatigue | 1.01 | 0.86 | 1.17 | 0.923 | 0.98 | 0.84 | 1.15 | 0.847 |
| Hallucinations | 1.44 | 0.62 | 2.89 | 0.349 | 1.40 | 0.60 | 2.88 | 0.388 |
| Headache | 0.88 | 0.72 | 1.08 | 0.231 | 0.82 | 0.66 | 1.00 | 0.058 |
| Language | 0.52 | 0.27 | 0.92 | 0.038 | 0.56 | 0.29 | 1.00 | 0.068 |
| Memory | 0.82 | 0.55 | 1.16 | 0.279 | 0.84 | 0.57 | 1.20 | 0.357 |
| Mood/anxiety | 0.67 | 0.56 | 0.80 | <0.001 | 0.63 | 0.52 | 0.75 | <0.001 |
| Sleep | 0.91 | 0.75 | 1.10 | 0.344 | 0.92 | 0.75 | 1.11 | 0.389 |

\* Models are adjusted for hospital type, age at admission, race, Hispanic ethnicity, public insurance, Charlson comorbidity index, ICU admission and mechanical ventilation.

**Figure 1.**
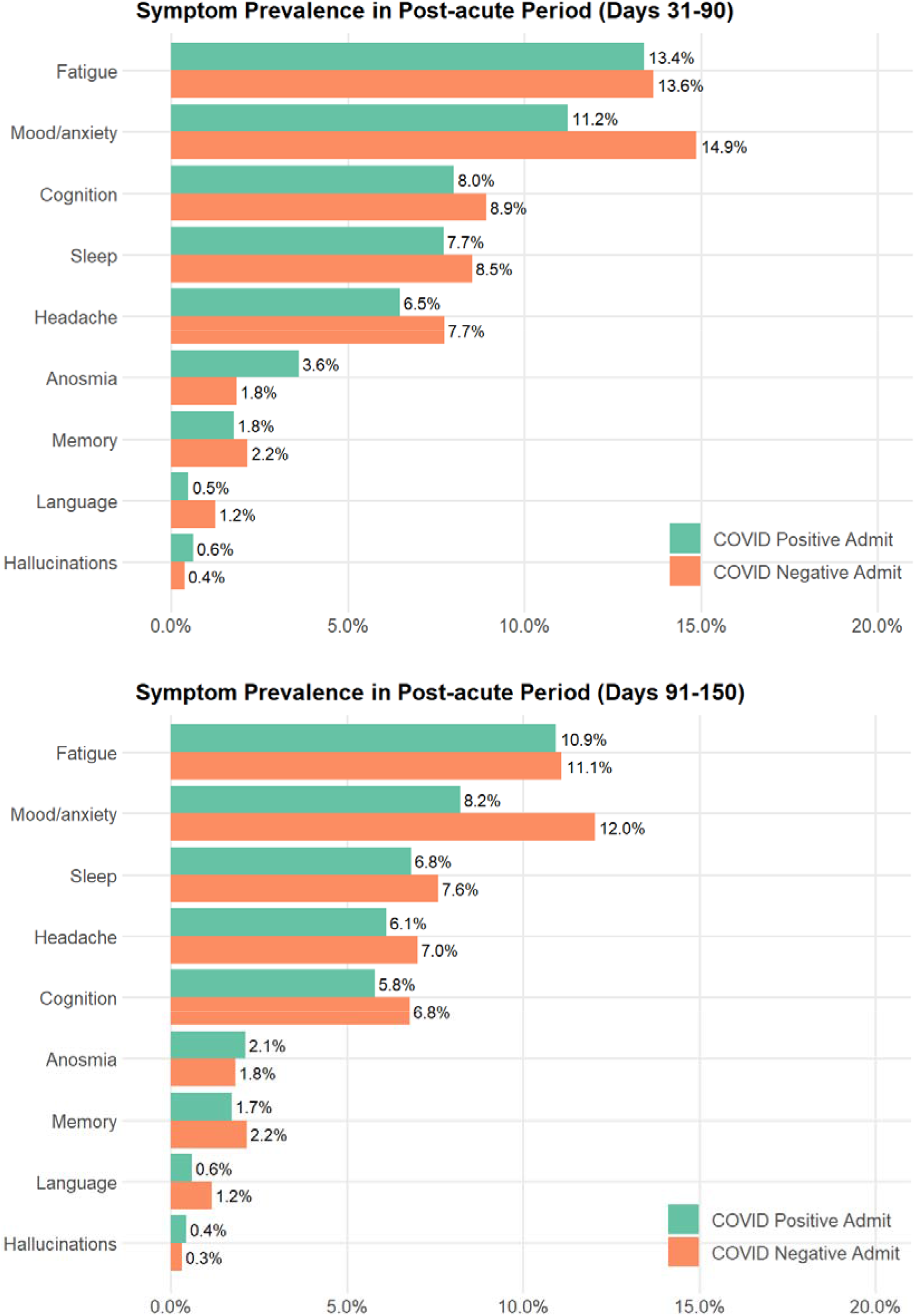
Frequency of new or persistent neuropsychiatric symptoms in COVID Positive vs COVID Negative admissions.

## Discussion

In this investigation of more than 40,000 hospitalized individuals over 12 months, including 6,619 individuals positive for SARS-CoV-2, we found that new-onset neuropsychiatric symptoms persisting at 30+ days occurred frequently but were not significantly more common among COVID-19 patients than individuals previously hospitalized for other reasons. Both language disturbance and affective symptoms were in fact significantly *less* common among those with prior COVID-19.

In general, the most commonly-observed symptoms in both follow-up periods are consistent with those estimated in a recent large meta-analysis incorporating data from 51 studies of variable follow-up duration[15]. The symptom prevalence we observe is approximately half of that reported in that meta-analysis; however, we report only incident symptoms, excluding those documented for a given individual prior to COVID-19. Notably, in the meta-analysis only 2 of 51 studies included control subjects, underscoring the need for more such comparisons.

On the other hand, our results are not fully consistent with those of a prior large study of post-acute sequelae among previously-hospitalized patients in the VA system[2]. In that study, greater rates of multiple neurologic and psychiatric diagnoses were identified compared to individuals hospitalized for influenza. Sensitivity analysis comparing that cohort of ∼13,000 to 900,000 hospitalized patients, using high-dimensionality matching, also found elevated rates of neuropsychiatric diagnoses among post-COVID-19 patients. Similarly, a claims-based study in ∼27,000 individuals diagnosed with COVID-19, utilizing multiple matched cohorts[3], identified elevated rates of amnesia, anxiety, and fatigue, among other symptoms. A more recent, large, diagnostic code-based study of similar design[4] likewise found elevated rates of anxiety, dementia, and psychotic disorders following acute illness compared to individuals diagnosed with influenza.

We note multiple important distinctions with the present work that may help explain this discordance. First, our study offers a more direct comparison to individuals hospitalized at the same time and in the same setting, avoiding the possibility that secular trends (for example, lesser acuity among hospitalized patients outside of COVID-19) could impact results. Second, we utilize natural language processing to examine symptoms rather than diagnostic codes alone, such that individual neurologic and psychiatric features may be captured even if not reflected in codes. In so doing, we may avoid Berkson’s bias[16], a form of collider bias in which post-COVID-19 individuals may, by receiving closer follow-up, be more likely to be given additional diagnoses. Indeed, a comparison of NLP and claims-based symptom descriptions with manual annotation of 3 sets of symptoms (Supplemental Table 3) indicates the extent to which claims codes alone may not adequately capture neuropsychiatric symptoms.

An earlier study of COVID-19 patients in Wuhan, China also found elevated rates of psychiatric symptoms at 6 months following hospitalization[6]. A key strength of that study was use of standardized rating scales and consistent follow-up interval, both limitations in interpreting our results. However, while that study supports the prevalence of such symptoms, it does not allow comparison to similar patients with difficult hospital courses attributable to other causes. That is, that study demonstrates that post-hospital course may be chronic in individuals with COVID-19, but not necessarily that this outcome is specific to COVID-19. Our results suggest that they may not be.

Among the more notable findings in the present study is the relative decrease in prevalence of mood and anxiety symptoms relative to non-COVID-19 patients. Among outpatients, these rates have been suggested to be elevated[2,4], consistent with a large survey-based study that also suggested differences in symptomatology[17]. The effects we observe may be specific to more severely ill individuals, such as those previously hospitalized. Some prior work may also reflect characteristics of particular subgroups, such as US military veterans (with likelihood of greater comorbidity[2]) or commercially-insured[4]; a strength of this study is the inclusion of the full spectrum of payers. Our discordant results also reflect the capture and documentation of symptoms, rather than diagnoses per se, in the present study – i.e., we are quantifying a different phenotype. At minimum, further investigation is needed to better understand these highly prevalent sequelae.

The strengths of the present study – namely, its use of natural language processing applied to large-scale electronic health records – also contribute to its limitations. First, follow-up is naturalistic, which may avoid the selection bias inherent in COVID-19 cohort studies, but also increases the risk that all participants are not uniformly observed. We attempted to minimize this risk by only including individuals with at least one observation during the follow-up period. We recognize that survival approaches could also be applied in this context, but elected to avoid this strategy as it would introduce a competing risk problem (i.e., individuals who die before entering a particular risk period could not experience a given symptom) and complicate interpretation of results and comparison to other studies. Similarly, we limited analysis to individuals with a hospital course of 30 days or less, recognizing that this will lead to undersampling of more severe illness course for both COVID positive and negative individuals. Second, we utilized natural language processing (as well as coded data reflecting symptoms) to identify individual neuropsychiatric characteristics, not diagnoses per se. This approach should be more sensitive to disease sequelae even if they do not rise to the level of a diagnosis; however, it is still less sensitive than systematic assessment at a fixed interval. Importantly, for the 3 symptoms that we are able to validate against gold standard, we show that the NLP-based approach differs in terms of sensitivity and specificity of the code-based approach, suggesting that neither approach alone is likely to be sufficient. Notably, numerous terms were omitted because of a lack of specificity (e.g., ‘flat’, as it relates to mood) – training classifiers for each individual symptom, if feasible, would undoubtedly improve symptom detection, at the cost of poorer generalizability. Finally, as we rely on two academic health systems in a single US region, the extent to which our results generalize to other regions or nations remains to be determined.

In aggregate, our results do not diminish the importance of further investigation of PASC as it relates to neuropsychiatric phenotypes. However, they suggest that strategies developed for rehabilitation of individuals with such symptoms following hospitalization, regardless of etiology, merit investigation in COVID-19. Undoubtedly the consequences of acute SARS-CoV-2 infection persist for a subset of individuals and have great capacity to diminish quality of life. Systematic investigations, including planned meta-cohorts in the US and elsewhere, will be critical in better defining these consequences. Our observation that they may not be specific to COVID-19 in no way detracts from the need to develop targeted interventions to address such symptoms – indeed, it highlights the potential utility of investigating a broad range of neuropsychiatric interventions[18] to address persistent cognitive and psychiatric symptoms[19].

## Data Availability

Data is not available for secondary use per IRB standards.

## Author Contributions

RP - conceived analysis, drafted and revised manuscript

VC - generated data set and analyzed data, revised manuscript

TM, JR, and JG – revised manuscript

## References

1. Bodro M, Compta Y, Sánchez-Valle R. Presentations and mechanisms of CNS disorders related to COVID-19. Neurol - Neuroimmunol Neuroinflammation. 2021;8.

2. Al-Aly Z, Xie Y, Bowe B. High-dimensional characterization of post-acute sequalae of COVID-19. Nature. 2021:1–8.

3. Daugherty SE, Guo Y, Heath K, Dasmariñas MC, Jubilo KG, Samranvedhya J, et al. Risk of clinical sequelae after the acute phase of SARS-CoV-2 infection: retrospective cohort study. BMJ. 2021;373:1098.

4. Taquet M, Geddes JR, Husain M, Luciano S, Harrison PJ. 6-month neurological and psychiatric outcomes in 236 379 survivors of COVID-19: a retrospective cohort study using electronic health records. Lancet Psychiatry. 2021;8:416–427.

5. Sudre CH, Murray B, Varsavsky T, Graham MS, Penfold RS, Bowyer RC, et al. Attributes and predictors of long COVID. Nat Med. 2021;27:626–631.

6. Huang C, Huang L, Wang Y, Li X, Ren L, Gu X, et al. 6-month consequences of COVID-19 in patients discharged from hospital: a cohort study. The Lancet. 2021;0.

7. Jaywant A, Vanderlind WM, Alexopoulos GS, Fridman CB, Perlis RH, Gunning FM. Frequency and profile of objective cognitive deficits in hospitalized patients recovering from COVID-19. Neuropsychopharmacol Off Publ Am Coll Neuropsychopharmacol. 2021. 15 February 2021. https://doi.org/10.1038/s41386-021-00978-8.

8. Nalichowski R, Keogh D, Chueh HC, Murphy SN. Calculating the benefits of a Research Patient Data Repository. AMIA Annu Symp Proc. 2006;2006:1044.

9. Murphy SN, Weber G, Mendis M, Gainer V, Chueh HC, Churchill S, et al. Serving the enterprise and beyond with informatics for integrating biology and the bedside (i2b2). J Am Med Inform Assoc JAMIA. 2010;17:124–130.

10. Charlson M, Szatrowski TP, Peterson J, Gold J. Validation of a combined comorbidity index. J Clin Epidemiol. 1994;47:1245–1251.

11. Barroilhet SA, Bieling AE, McCoy TH, Perlis RH. Association between DSM-5 and ICD-11 personality dimensional traits in a general medical cohort and readmission and mortality. Gen Hosp Psychiatry. 2020;64:63–67.

12. McCoy TH, Yu S, Hart KL, Castro VM, Brown HE, Rosenquist JN, et al. High Throughput Phenotyping for Dimensional Psychopathology in Electronic Health Records. Biol Psychiatry. 2018;83:997–1004.

13. Menni C, Valdes AM, Freidin MB, Sudre CH, Nguyen LH, Drew DA, et al. Real-time tracking of self-reported symptoms to predict potential COVID-19. Nat Med. 2020;26:1037–1040.

14. R Core Team. R: A Language and Environment for Statistical Computing. Vienna, Austria: R Foundation for Statistical Computing; 2019.

15. Badenoch JB, Rengasamy ER, Watson CJ, Jansen K, Chakraborty S, Sundaram RD, et al. Persistent neuropsychiatric symptoms after COVID-19: a systematic review and meta-analysis. MedRxiv. 2021:2021.04.30.21256413.

16. Westreich D. Berkson’s bias, selection bias, and missing data. Epidemiol Camb Mass. 2012;23:159–164.

17. Perlis RH, Santillana M, Ognyanova K, Green J, Druckman J, Lazer D, et al. Comparison of post-COVID depression and major depressive disorder. MedRxiv. 2021:2021.03.26.21254425.

18. Balas MC, Vasilevskis EE, Olsen KM, Schmid KK, Shostrom V, Cohen MZ, et al. Effectiveness and safety of the awakening and breathing coordination, delirium monitoring/management, and early exercise/mobility bundle. Crit Care Med. 2014;42:1024–1036.

19. ServickApr. 27 K, 2021, Pm 2:40. COVID-19 ‘brain fog’ inspires search for causes and treatments. Sci AAAS. 2021. https://www.sciencemag.org/news/2021/04/covid-19-brain-fog-inspires-search-causes-and-treatments. Accessed 30 April 2021.

