## Supplemental Material for "Case-control study of neuropsychiatric symptoms following COVID-19 hospitalization in 2 academic health systems"

**Supplemental Table 1.** Symptom NLP Search Term

| **Symptom** | **Search term** |
| --- | --- |
| Memory | "Memory impairment" |
| Memory | "Memory loss" |
| Memory | "Getting lost" |
| Memory | "Difficulty remembering" |
| Memory | "Decreased memory" |
| Memory | "Loss of memory" |
| Cognition | "Poor Cognition" |
| Cognition | "Impaired cognition" |
| Cognition | "Cognitive impairment" |
| Cognition | "Brain fog" |
| Cognition | "Impaired Concentration" |
| Cognition | "Trouble with concentration" |
| Cognition | "Trouble Concentrating" |
| Cognition | "Difficulty concentrating" |
| Cognition | "Loss concentration" |
| Cognition | "Difficulty focusing" |
| Cognition | "Trouble focusing" |
| Cognition | "Loss focus" |
| Cognition | "Executive Functioning" |
| Cognition | "Difficulty planning" |
| Cognition | "Difficulty organizing" |
| Cognition | "Trouble planning" |
| Cognition | "Trouble organizing" |
| Cognition | "trouble Decision-making" |
| Cognition | "difficult Decision making" |
| Cognition | "difficulty making decisions" |
| Cognition | Confusion |
| Cognition | Confused |
| Cognition | "Slowed thoughts" |
| Cognition | "Slowed thinking" |
| Language | "Trouble speaking" |
| Language | "Trouble finding words" |
| Language | "Trouble word finding" |
| Language | "Difficulty speaking" |
| Language | "Difficulty finding words" |
| Language | "Difficulty word finding" |
| Language | "Can’t find words" |
| Language | "Can’t remember names" |
| Language | "Difficulty names" |
| Language | Agnosia |
| Language | "Poor word retrieval" |
| Language | "Difficulty word retrieval" |
| Language | "Difficulty writing" |
| Language | "Trouble Writing" |
| Language | "Trouble Speaking" |
| Language | "Trouble Communicating" |
| Sleep | "Broken sleep" |
| Sleep | "Restless sleep" |
| Sleep | "Decreased sleep" |
| Sleep | "Increased sleep" |
| Sleep | "Poor sleep" |
| Sleep | "Disrupted sleep" |
| Sleep | Insomnia |
| Sleep | "Restless legs" |
| Sleep | "Sleep apnea" |
| Sleep | "Vivid dreams" |
| Sleep | Nightmares |
| Headache | Headache |
| Headache | Migraine |
| Mood/anxiety | Anxiety |
| Mood/anxiety | Anxious |
| Mood/anxiety | Irritable |
| Mood/anxiety | Irritability |
| Mood/anxiety | Anger |
| Mood/anxiety | Angry |
| Mood/anxiety | Depressed |
| Mood/anxiety | Depression |
| Mood/anxiety | Apathy |
| Mood/anxiety | "Mood swings" |
| Mood/anxiety | Moody |
| Mood/anxiety | "Mood lability" |
| Mood/anxiety | Suicidal |
| Mood/anxiety | Suicidality |
| Mood/anxiety | Suicide |
| Mood/anxiety | Mania |
| Mood/anxiety | Manic |
| Mood/anxiety | Hypomania |
| Mood/anxiety | Hypomanic |
| Anosmia | "Loss Smell" |
| Anosmia | "Loss Taste" |
| Anosmia | "Change smell" |
| Anosmia | "Change taste" |
| Anosmia | "Loss olfaction" |
| Anosmia | "Change olfaction" |
| Anosmia | Ageusia |
| Anosmia | Dysgeusia |
| Anosmia | Anosmia |
| Anosmia | Dysosmia |
| Hallucinations | "Visual hallucination" |
| Hallucinations | "Auditory hallucination" |
| Hallucinations | "Hearing things" |
| Fatigue | "Poor Energy" |
| Fatigue | "Loss of energy" |
| Fatigue | Tired |
| Fatigue | Fatigue |
| Fatigue | Malaise |
| Fatigue | Tiredness |
| Fatigue | Napping |
| Fatigue | Exhaustion |
| Fatigue | Exhausted |
| Fatigue | "Decreased activity" |
| Fatigue | "Decrease activity" |
| Fatigue | "Reduced activity" |

**Supplemental Table 2**. Symptom ICD Code Definitions

| **symptom** | **ICD10CD** | **ICD10CD description** |
| --- | --- | --- |
| Anosmia | R43.0 | Anosmia |
| Anosmia | R43.1 | Parosmia |
| Anosmia | R43.2 | Parageusia |
| Anosmia | R43.8 | Other disturbances of smell and taste |
| Anosmia | R43.9 | Unspecified disturbances of smell and taste |
| Cognition | R40.4 | Transient alteration of awareness |
| Cognition | R41.0 | Disorientation, unspecified |
| Cognition | R41.82 | Altered mental status, unspecified |
| Cognition | R41.840 | Attention and concentration deficit |
| Cognition | R41.841 | Cognitive communication deficit |
| Cognition | R41.842 | Visuospatial deficit |
| Cognition | R41.843 | Psychomotor deficit |
| Cognition | R41.844 | Frontal lobe and executive function deficit |
| Cognition | R41.89 | Other symptoms and signs involving cognitive functions and awareness |
| Cognition | R41.9 | Unspecified symptoms and signs involving cognitive functions and awareness |
| Fatigue | R53.81 | Other malaise |
| Fatigue | R53.82 | Chronic fatigue, unspecified |
| Fatigue | R53.83 | Other fatigue |
| Hallucinations | R44.0 | Auditory hallucinations |
| Hallucinations | R44.1 | Visual hallucinations |
| Hallucinations | R44.2 | Other hallucinations |
| Hallucinations | R44.3 | Hallucinations, unspecified |
| Language | R47.01 | Aphasia |
| Language | R47.02 | Dysphasia |
| Language | R47.1 | Dysarthria and anarthria |
| Language | R47.81 | Slurred speech |
| Language | R47.82 | Fluency disorder in conditions classified elsewhere |
| Language | R47.89 | Other speech disturbances |
| Language | R47.9 | Unspecified speech disturbances |
| Memory | R41.1 | Anterograde amnesia |
| Memory | R41.2 | Retrograde amnesia |
| Memory | R41.3 | Other amnesia |
| Mood/anxiety | R45.1 | Restlessness and agitation |
| Mood/anxiety | R45.3 | Demoralization and apathy |
| Mood/anxiety | R45.4 | Irritability and anger |
| Mood/anxiety | R45.6 | Violent behavior |
| Mood/anxiety | R45.7 | State of emotional shock and stress, unspecified |
| Mood/anxiety | R45.81 | Low self-esteem |
| Mood/anxiety | R45.84 | Anhedonia |
| Mood/anxiety | R45.850 | Homicidal ideations |
| Mood/anxiety | R45.851 | Suicidal ideations |
| Mood/anxiety | R45.86 | Emotional lability |
| Mood/anxiety | R45.87 | Impulsiveness |
| Mood/anxiety | R45.89 | Other symptoms and signs involving emotional state |
| Sleep | R40.0 | Somnolence |

**Supplemental Table 3.** Evaluation of 3 selected symptoms, defined by ICD10 code or NLP tokens in 909 admissions manually abstracted as part of the MGH COVID Registry.

| **Symptom** | **Source** | **Balanced accuracy** | **Specificity** | **Sensitivity** | **PPV** | **NPV** |
| --- | --- | --- | --- | --- | --- | --- |
| anosmia | ICD10 | 0.503 | 0.999 | 0.008 | 0.500 | 0.863 |
| anosmia | NLP | 0.745 | 0.810 | 0.680 | 0.363 | 0.941 |
| headache | ICD10 | 0.553 | 0.941 | 0.165 | 0.403 | 0.824 |
| headache | NLP | 0.643 | 0.457 | 0.830 | 0.268 | 0.918 |
| fatigue | ICD10 | 0.518 | 0.891 | 0.144 | 0.343 | 0.725 |
| fatigue | NLP | 0.558 | 0.201 | 0.914 | 0.311 | 0.856 |

**Supplemental Figure 1.** Symptom Prevalence in the Acute Period (-14 to 30 days from COVID index date)

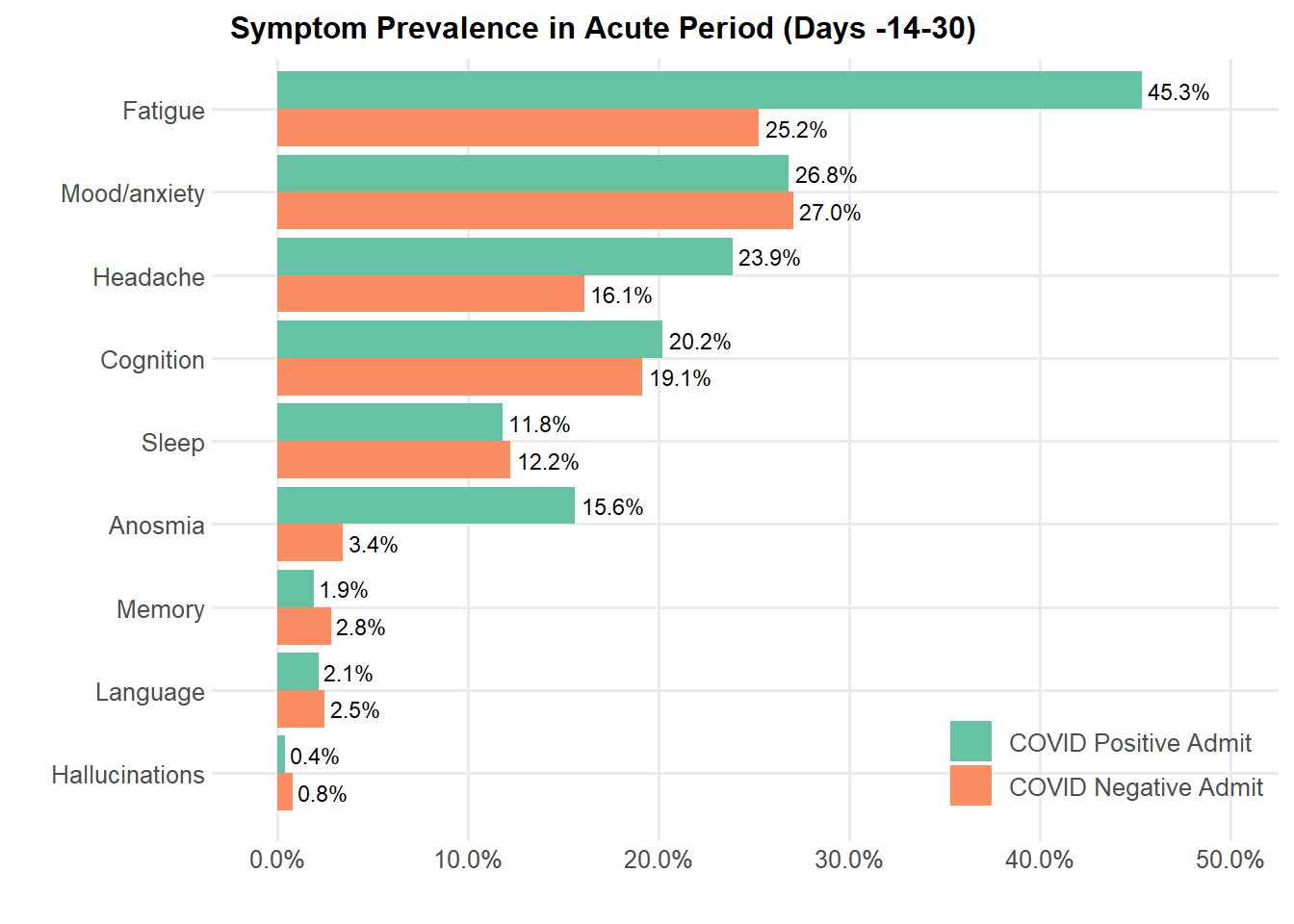
